# Two waves of the pertussis epidemic in England in 2023 and 2024

**DOI:** 10.1101/2024.04.28.24306493

**Authors:** Igor Nesteruk

## Abstract

The resurgence of pertussis (whooping cough) becomes a serious problem in many countries including the UK. Differentiation of the accumulated monthly numbers of pertussis cases registered in England in 2023 and 2024 revealed two waves of the epidemic before and after October 2023. Identification of parameters of SIR (susceptible-infectious-removed) model allowed calculating the numbers of infectious persons and reproduction rates. The accumulated and daily numbers of cases and the duration of the first wave were predicted. If the influence of second wave will be not very significant, the new cases will stop to appear in the end of August 2025 after reaching the figure of 5.8 thousand. The maximum of average daily numbers of new cases is expected to be around 51 on 9-10 May 2024. Since the effective reproduction number is very close to its critical value 1.0, the probably of new outbreaks is very high. May be the, increase of percentage of vaccinated people could decrease this probability.

## Introduction

The resurgence of pertussis (whooping cough) increases the risk of infant fatality and became a serious problem in many countries including the developed ones [1-16]. Many researchers explain the increasing numbers of cases by decreasing the level of vaccinations and propose maintain the immunization at the level more than 90% worldwide [4].

Some mathematical studies are trying to improve the control of the pertussis outbreaks [2, 5]. In particular, the age structure of the population has been taken into account [2, 5]. A modified SIR (susceptible-infectious-removed) model was used in [5] and some results of calculations are presented. Nevertheless, we have not found any solutions of inverse problems for pertussis outbreaks, i.e. identifications of the model parameters or the reproduction numbers with the use of real datasets as it was done for the COVID-19 pandemic dynamics [17-23].

In this study we will use the accumulated numbers of pertussis cases registered in England in 2023 and 2024 (14 months of a recent outbreak, [1]), SIR model and the method of identification of its parameters, proposed in [24] and successfully used in [17-19, 25, 26]. The numbers of infectious person and the reproduction rates will be calculated. We will try to predict the accumulated numbers of cases and averaged daily numbers of new cases and the duration of the recent pertussis outbreak in England.

## Materials and Methods

We will use the accumulated numbers *V*_*j*_ of laboratory-confirmed pertussis cases in England, [1] (shown in Table 1). Since only monthly data is available (with different numbers of days), we have calculated the accumulated numbers of days *t*_*j*_ starting with 1 January 2023.

**Table 1.** Accumulated numbers of confirmed pertussis cases in England in 2023 and 2024 and estimations of the average daily numbers of cases and its derivative.

| Months, starting with January 2023, [1] | Days, starting with 1 January 2023, $t_j$ | Accumulated numbers of cases, $V_j$, [1] | Calculated daily values of $dV/dt$, eq. (1) | Calculated daily values of $d^2V/dt^2$, eq. (2) |
| --- | --- | --- | --- | --- |
| 1 | 31 | 9 | - | - |
| 2 | 59 | 18 | 0.35426 | 0.0023453 |
| 3 | 90 | 30 | 0.52688 | 0.0090184 |
| 4 | 120 | 50 | 0.86559 | 0.0132616 |
| 5 | 151 | 83 | 1.41559 | 0.0226500 |
| 6 | 181 | 136 | 2.04462 | 0.0185305 |
| 7 | 212 | 208 | 2.66129 | 0.0218522 |
| 8 | 243 | 301 | 3.20 | 0.0129032 |
| 9 | 273 | 403 | 3.34516 | -0.0036559 |
| 10 | 304 | 505 | 3.47849 | 0.0121401 |
| 11 | 334 | 615 | 5.75269 | 0.1390681 |
| 12 | 365 | 858 | 12.87096 | 0.3246618 |
| 13 | 396 | 1413 | 24.69299 | 0.4380494 |
| 14 | 425 | 2326 | - | - |

Changes in social behavior, quarantine measures; appearing new strains, etc. may change a pandemic dynamics. To detect the corresponding new waves, first and second derivatives of the accumulate numbers of cases could be useful [17, 18]. Since the accumulated numbers of cases *V*_*j*_ are given for every month, there is no need to smooth these values (in comparison with the COVID-19 pandemic, where the 7-days smoothing was used for very random daily datasets, [17, 18]). To estimate the average daily numbers of cases and the rate of their change the first and second derivatives can be estimated with the use of simple formulae:

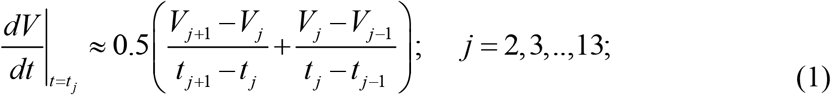

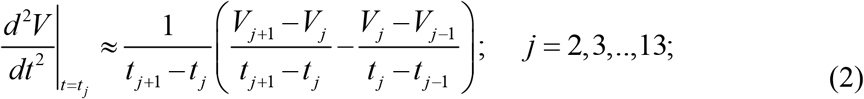

The corresponding monthly characteristics can be obtained by substituting *t*_*j*_ by

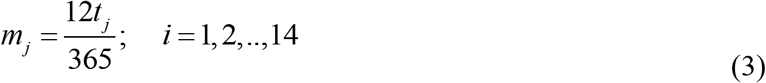

in formulae (1) and (2). Eq. (3) takes into account different lenghs of the months.

For every epidemic wave *i*, the generalized SIR model can be applied, which relates numbers of susceptible *S(t)*, infectious *I(t)* and removed persons *R(t)* versus time *t*, [17, 19]:

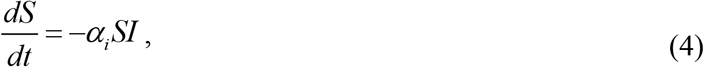

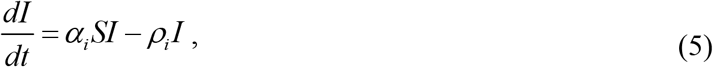

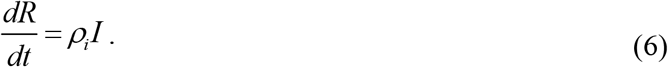

Infection and removal rates (*α*_*i*_ and *ρ*_*i*_) are supposed to be constant for every epidemic wave, i.e. for the time periods: 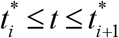. The inverse values 1/ *ρ*_*i*_ are the estimations of the average time of spreading infection *τ*_*i*_ (or the generation time [20]) during *i-th* epidemic wave

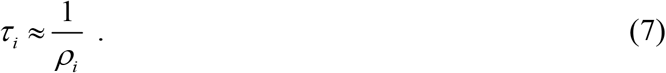

Summarizing eqs. (4)-(6) yields zero value of the derivative *d* (*S* + *I* + *R*) / *dt*. Then the sum:

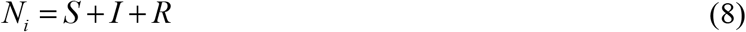

must be constant for every epidemic wave. We consider the value *N*_*i*_ to be an unknown parameter of the SIR model corresponding to the *i-th* wave, which must be estimated by observations. There is no need to assume that this constant equals the known volume of population and to reduce the problem to a 2-dimensional one. Many researchers (see, e.g., [5]) use this additional unrealistic condition, which means that before the outbreak all people are susceptible. However, many people are protected by their immunity, distance, lockdowns, etc. In the case of pertussis, the rejection of total susceptibility assumption is especially important, since the disease mainly affects children, most of who are vaccinated. Even in the case of respiratory COVID-19 infection, estimates of the initial number of susceptible people (before the outbreak) in China yielded values between 91 and 138 thousand, (0.006% - 0.01% of the population), [17].

The initial conditions for the set of equations (4)–(6) at the beginning of every epidemic wave 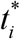 can be written as follows

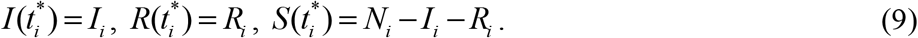

The exact solution of the set of non-linear differential equations (4)-(6) can be obtained using the function

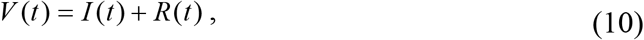

which corresponds to the number of victims or the cumulative numbers of cases over time *t* and has the following form [17, 19]

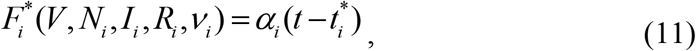

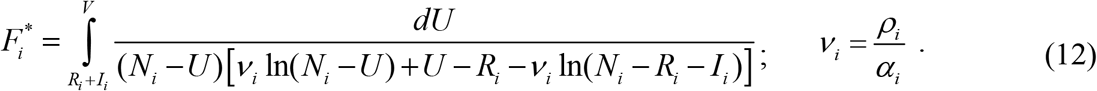

Thus, for every set of parameters 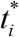 and a fixed value of *V*, integral (12) can be calculated and a corresponding moment of time can be determined from (9). The *S(t), I(t)* and *R(t)* values can be calculated with the use of the following equations [20, 21]:

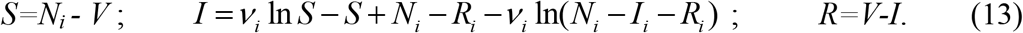

The derivative *dV/dt* yields the estimate of the average daily number of new cases and with the use of eqs. (5) and (6) can be written as follows:

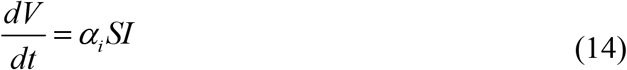

Using (4)-(6), (10), and(14) the second derivative can be expressed as follows, [17]:

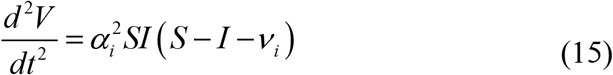

Zero value of the second derivative (15) corresponds to the maximum of theoretical estimation (14) of the average daily (or monthly) numbers of new cases. Corresponding values of susceptible persons *S*_*s*_ and moment of time *t*_*s*_ can be calculated with the use of formulas available in [17]. At *t>t*_*s*_ the second derivative (15) cannot be positive, since *S(t)* diminishes monotonously (see (4)). Therefore, a change in the sign of the second derivative from negative to positive reflect the beginning of a new epidemic wave [17, 18].

The effective reproduction number *R*_*t*_*(t)* shows the average number of people infected by one person and can be calculated with the use of different approaches, [21-23, 25, 27, 28]. The generalized SIR model also allows estimating the reproduction numbers for each epidemic wave with the use of the formula, [26]:

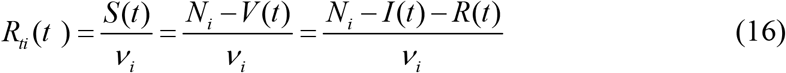

Eqs. (15) and (16) illustrate that at the moment *t*_*s*_ of maximum daily cases (14) the reproduction number is still higher than the critical value 1.0 (i.e. *R*_*ti*_ (*t*_*s*_) = 1+ *I* (*t*_*s*_) /*ν*_*i*_), but diminishes monotonously and approaches

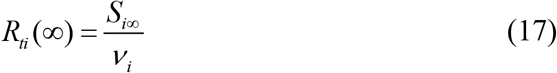

for every epidemic wave *i*, where the number of susceptible persons at infinity *S*_*i*∞_ can be found from the non-linear equation, [17, 26]:

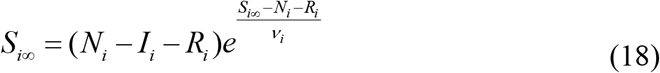

To estimate the final day of the *i-th* epidemic wave, we can use the condition:

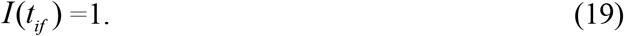

which means that at *t* > *t*_*if*_, less than one person still spreads the infection [17, 26]. Similar condition can be used to calculate the moment *t*_*i*0_, when a “zero” patient has appeared [26]:

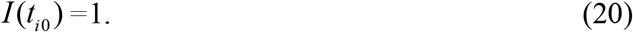

Usually new epidemic waves are connected with new pathogen strains (see e.g. [29, 30]). Then the value *t*_*i*0_ could indicate the time when the new variant (that caused the *i-th* epidemic wave) started to circulate in a population.

In the case of the first epidemic wave, we can use the values *I*_1_ =1; *R*_1_ =0. Then 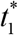 can be treated as the moment of appearance of “zero” patient. Immediately after an epidemic outbreak, *N*_1_ >> *V* ≥ 1, and integral (12) can be simplified as follows 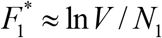. Then eq. (11) yields

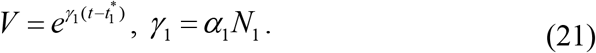

I.e., the exponential growth of the accumulated numbers of cases, the daily numbers of cases *dV/dt*, and the second derivative (15) is typical for the initial stages of epidemics. Knowing the value of the parameter *γ*_1_, the time of cases duplication can be calculated as follows [17, 26]:

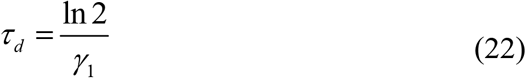

Different procedures for parameter identification can be found in [17-19, 24, 26], but all of them use the accumulated numbers of cases *V*_*j*_ registered during some period of time. Then the corresponding values 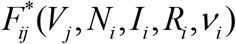 can be calculated with the use of (12). Due to the linear relationship (11) the linear regression between 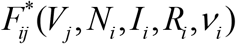 and corresponding *t*_*j*_ values can be used to find the correlation coefficient *r*, best fitting lines, and parameters *α*_*i*_ and 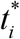, [31]. Optimal values of the parameters have to ensure maximal values of the correlation coefficient or Fisher function:

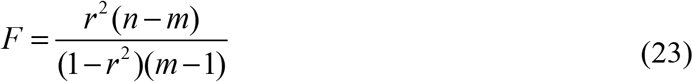

where *n* is the number of observations; *m*=2 is the number of parameters in the regression equation, [31].

In particular, for the first epidemic wave there are only two independent parameters *N*_*i*_ and *ν*_*i*_. After finding their optimal values corresponding to maximum of *r*, the theoretical SIR curves can be calculated with the use of (11) - (13). These dependences correspond to best fitting with the random results of observations and can be used for prediction the future epidemic dynamics (in particular, to estimate the numbers of infectious persons and reproduction rates). These approach was proposed in [16] and successfully used for simulations of a mysterious children disease in Ukrainian city Chernivtsi [24] and different waves of the COVID-19 pandemic [17-19, 25, 26].

## Results and Discussion

The registered accumulated numbers of pertussis cases [1] and calculated values of monthly derivatives (according to eqs. (1) –(3), the daily characteristics shown in Table 1 have to be increased according to eq. (3)) are shown in Fig.1. The use of logarithmic scale allows detecting the periods of exponential grows. After the outbreak, corresponding values follow the straight lines (according to eq. (21)). Very good coincidences are visible for *V*_*j*_ values (blue) between February and July 2023; for the first derivative (eq. (1), black) - between March and June 2023; and for the second derivative (eq. (2), red) - between March and May 2023.

To estimate the value of the parameter *γ*_1_ let us use eq. (21) and values of *V*_*j*_ and *t*_*j*_ (*j*=2 and 7) listed in Table 1.

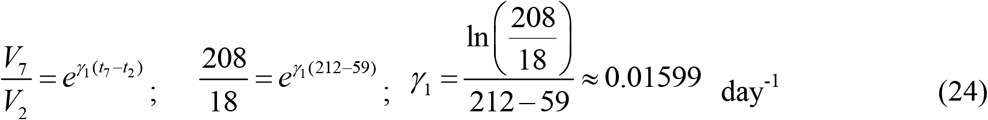

With the use of (22) and (24) the time of cases duplication can be estimated as 43.34 days. The bacterial pertussis infection spreads much slower, than respiratory ones (e.g., COVID-19 cases duplicated in 2.3 - 3.7 days in February-March 2020, [17, 32]).

After July 2023, the pertussis epidemic dynamics demonstrated a stabilization. The monthly numbers of new cases were almost constant in August, September and October (see Table 1 and the black line in Fig.1), but after October 2023 we see the increasing trend again (see Table 1 and the black line in Fig.1).

The second derivative has changed its sign from negative to positive in October 2023 (see Table 1 and the red line in Fig. 1). It means that a new epidemic wave has started and we cannot use all available *V*_*j*_ values to calculate the optimal values of SIR model. Our attempts to use the algorithm presented in previous Section for complete dataset (*n*=14), were not successful. Nevertheless, with the use of first nine *V*_*j*_ values, the maximum of the correlation coefficient was isolated.

**Fig. 1.**
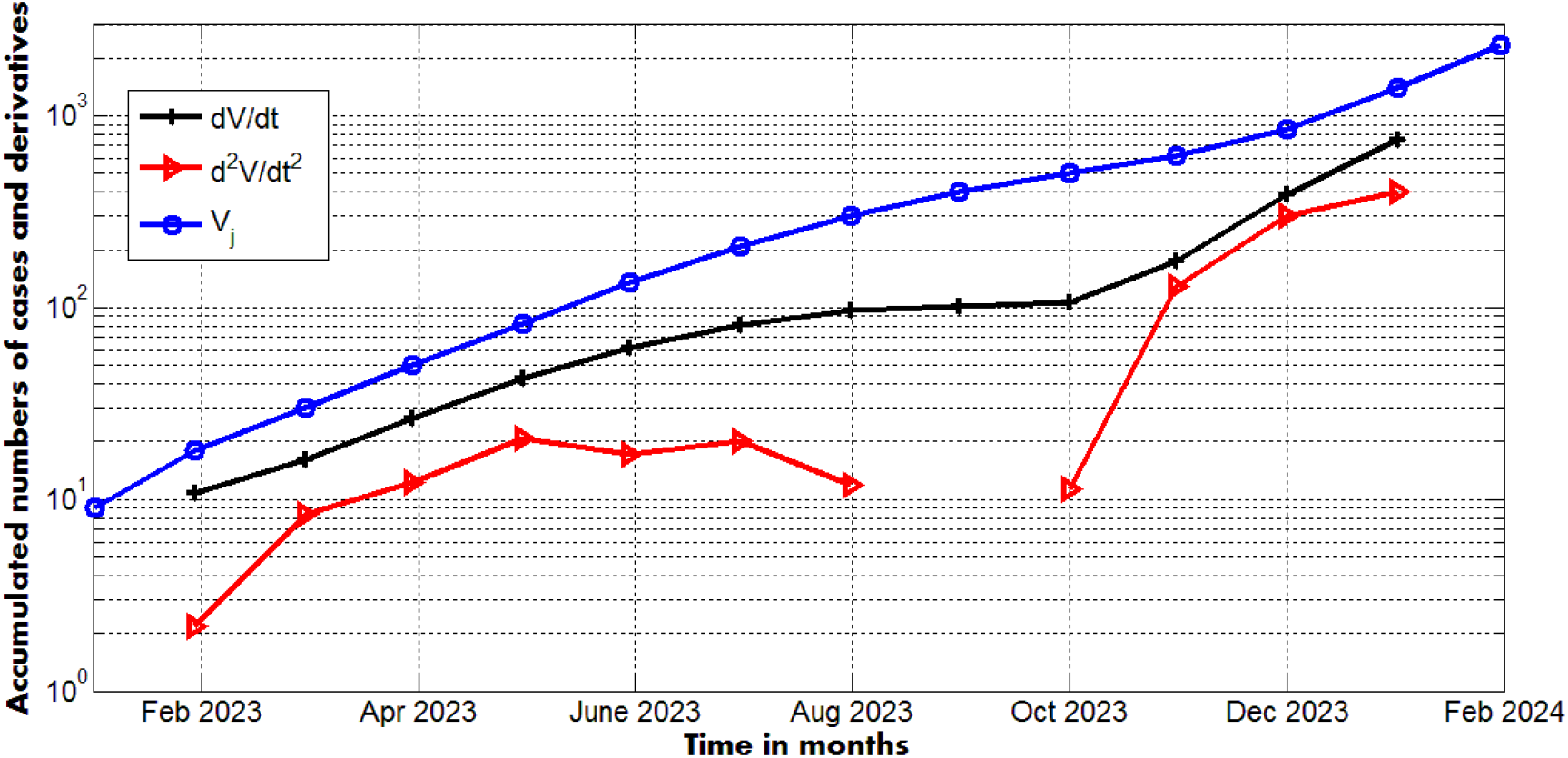
Accumulated numbers of pertussis cases in England ([1], blue), average monthly number of new cases *dV/dt* (black, eqs. (1) and (3)) and its derivative *d*^*2*^*V/dt*^*2*^ ((eqs. 2) and (3)).

Fig. 2 represents the calculated SIR curves (4)-(6), the first and second derivatives (14) and (15), and reproduction number (16). The optimal values of parameters (corresponding to the maximal value of correlation coefficient *r*=0.998545303649538) are:

**Fig. 2.**
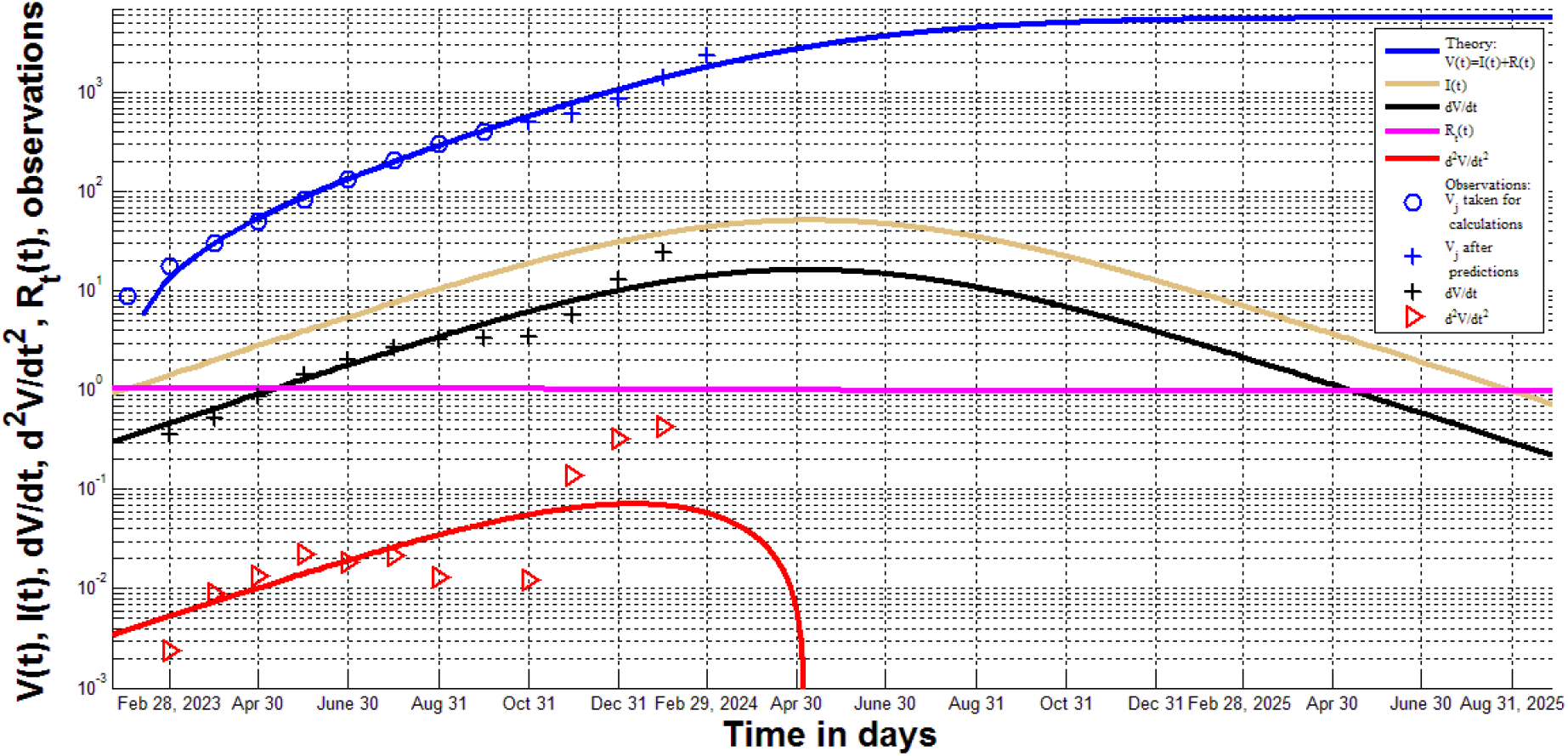
Results of calculations with the use SIR model (curves) and observations (markers). Accumulated numbers of pertussis cases (blue; curve corresponds to *V=I+R*, eq. (10)); “circles” - to the values taken for identification of SIR model parameters; “crosses” represent observations after the prediction). The average daily numbers of new cases (black; curve - eq. (14); “crosses” – eq. (1), Table 1). The second derivative *d*^*2*^*V/dt*^*2*^ (red; curve - eq. (15); “triangles” –eq. (2), Table 1). The magenta line represents the reproduction number (eq. (16)). The brown line show the numbers of infectious persons *I(t)* (eq. (13)).

N_1_=84204.5032875141;

*ν*_1_ =81306.0503840472;

*α*_1_ =3.89331528144046e-06 [day]^-1^;

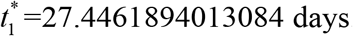.

Very high values of the correlation coefficient and the Fisher function (*F*=2401, eq. (23)) demonstrate that the linear dependence (11) is supported by the results of observations at the confidence level higher than 0.001 (corresponding critical value *F*_*c*_(7,1)=29.2, [33]).

The average time of spreading the infection (according to eq.(7)) *τ*_1_ ≈ 3.16 days. Since this value is not a duration of the illness, but reflects the speed and quality of isolation of invectives, it could be similar for different diseases in one country. In particular, during the first COVID-19 epidemic wave in the UK, *τ*_1_ was estimated as 3.03, [17]. The mean UK household generation time was estimated as 3.2 days for the Delta variant and 4.5 days for the Alpha variant [20]. The value of 21 days used for estimations in [5] looks unrealistic (at least for England).

The value 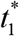 demonstrates that the first pertussis patient has started to infect between January 27 and 28, 2023. The smaller value of *V*_*1*_ (in comparison with the exponential trend for *V*_*2*_ *–V*_*7*_) probably reflects the fact that outbreak occurred not in the beginning of January 2023. It would be interesting to compare this result with the observations.

Fig. 2 shows rather good coincidence between the theoretical predictions for the accumulated numbers of cases (the blue line, eq. (10)) and results of observations *V*_*j*_ after the prediction (blue crosses). There are more discrepancies between the theoretical estimation of the daily numbers of new cases (eq. (14), the black curve) and results of calculations listed in Table 1 (black “crosses”). For the second derivative, the results of observations (Table 1, red “triangles”) significantly deviates from the theoretical red curve (eq. (15)). In particular, the negative value of *d*^*2*^*V/dt*^*2*^ (not shown in Figs. 1 and 2) was registered in September 2023 (see Table 1).

The final number of susceptible person for the first epidemic wave *S*_1∞_ = 78,448 (eq. (18)) and the final number of the reproduction number is 0.9649 (eq. (17)), while the initial values were 84,205 and 1.0357, respectively. High *S*_1∞_ values and reproduction rates, which are close to the critical value 1.0 (see the magenta curve in Fig. 2), show that the probabilities of a new epidemic wave or a new outbreak (after the final moment *t*^1 *f*^ (eq. (19)) are very high. The reproduction numbers in September were still supercritical (varied form 1.0320 to 1.0306). Most likely, we are already observing a new wave after September 2023. If the influence of this wave will be not very significant, we could expect the final number of accumulated cases *V*_1∞_ = *N*_1_ − *S*_1∞_ around 5,756 (see the blue line in Fig. 2). Observations during next months will demonstrate the influence of the second wave on the accuracy of long-term predictions.

The presented SIR simulation of the first wave demonstrate rather low numbers of infectious persons (with the maximum of 51 around 9-10 May 2024 and less than 1.0 value after the end of August 2025, see the brown curve). The maximum of the average new daily cases (*dV/dt*, the black line) is expected around 6-7 May 2024. Nevertheless, the second and probably other waves can change these predictions. Unfortunately, we have only 5 data points corresponding to the second wave (*j* = 10 - 14). The limited number of observations makes SIR simulations of the second wave impossible, but they can be done later or with the use of recent weekly information about the accumulated numbers of cases.

## Conclusions

Two waves of the pertussis outbreak in England in 2023 and 2024 were revealed with the use of differentiation of the accumulated numbers of cases. SIR model, the algorithm of its parameter identification, and data corresponding to the period January-September 2023 were used to calculate and predict the accumulated and daily numbers of cases, numbers if infectious persons, and effective reproduction number.

If the influence of second wave (started in October 2024) will be not very significant, the new cases will stop to appear after the end of August 2025 after reaching the figure of 5.8 thousand. The maximum of average daily numbers of new cases is expected to occur around 6-7 May 2024.

The calculated values of the effective reproduction number are very close to its critical value 1.0. This fact and rather high final numbers of the susceptible persons increase the probability of new outbreaks. May be the, increase of percentage of vaccinated people could decrease this probability.

## Clarification point

No humans or human data was used during this study

## Data availability

All data generated or analyzed during this study are included in this text.

## Acknowledgement

The study was supported by INI-LMS Solidarity Programme at the University of Warwick, UK. The author is grateful to Robin Thompson, Matt Keeling, Paul Brown, and Oleksii Rodionov for their support and providing very useful information.

